# Female genital cutting and maternal attitudes about it: Testing a cultural disempowerment hypothesis across two large multi-country samples

**DOI:** 10.64898/2026.04.14.26350909

**Authors:** Paul S. Strand, Justin C. Trang

## Abstract

Female genital cutting (FGC) refers to the cultural practice of cutting or mutilating the genitalia of girls and women and is identified within global health and human rights discourse as a contributor to gender inequality and female disempowerment. The cultural repression hypothesis proposes that societies in which FGC is more prevalent may be characterized by social and cultural conditions that reduce women’s decision-making authority and autonomy than societies in which FGC is less prevalent. Yet anthropological research has questioned whether this interpretation adequately reflects how FGC is organized and maintained within practicing communities. Across two studies with 176,728 participants from 15 African and Asian countries, we examined the association between mothers’ attitudes about FGC and daughters FGC status. We hypothesized that this association would vary as a function of regional FGC prevalence, revealing greater association strength in societies in which the practice is less common. Consistent with this prediction, multilevel logistic regression models show that maternal attitudes strongly predict daughter circumcision status and that the association between maternal attitudes and daughter outcomes is stronger in geographic regions in which FGC is less prevalent. Findings replicated across two large, multi-country samples and was robust to daughter age differences across regions, mother education, and family income. They suggest that high FGC prevalence may reflect social dynamics that are restrictive of mother agency regarding daughter FGC outcomes. Policy implications of the findings are discussed.

---

Female genital cutting (FGC) refers to a set of practices involving the cutting and mutiliation of female genitalia that occur in numerous societies across Africa, the Middle East, parts of Asia, and their diasporas^1–3^. International policy and public health discourse commonly interpret the practice as a manifestation of gender inequality and structural constraints on women’s autonomy and agency^4–7^. Within this framework, the persistence of FGC is frequently attributed to restrictive social arrangements in which women have limited influence over decisions concerning their own bodies and those of their daughters^8–10^.

These assumptions have shaped international policy responses, including legal prohibitions, public health campaigns, and educational interventions aimed at promoting abandonment of the practice^6, 11–12^. Implicit in many such initiatives is the expectation that reductions in FGC prevalence will accompany welcomed increases in women’s autonomy, decision-making authority, and health^13^.

However, anthropological and cross-cultural scholarship has raised questions about whether this interpretation accurately reflects the social organization of FGC in practicing communities^14–19^. Ethnographic studies describe the practice as embedded within complex systems of kinship, social status, and intergenerational female networks that facilitate rather than proscribe equality and female empowerment^20–22^. These observations have led some scholars to argue that prevailing policy frameworks may oversimplify or mischaracterize the social dynamics surrounding FGC^23^. Rather than being imposed by male-dominated authority structures, the practice may in many contexts exist within organizational structures in which women themselves are primary participants^19, 23^; and may reflect gender equality given that FGC occurs only in societies that practice male circumcision^3^. Critics have further suggested that international policy discourse may sometimes interpret FGC primarily through external normative frameworks that do not fully capture local understandings of gender, social status, or family obligation^18, 22, 24^. According to this perspective, understanding the persistence of the practice requires examination of the social institutions and decision-making processes within communities themselves, to test assumptions across contexts that differ in the prevalence of FGC and other traditionalist cultural practices.

One way of examining the social conditions under which FGC is reproduced within families is to assess the correspondence between parental attitudes and offspring outcomes. If women’s preferences are substantially constrained by family or community expectations, maternal attitudes toward the practice may have less correspondence with whether daughters undergo FGC. This possibility may be particularly relevant in contexts where the practice is highly prevalent, where strong social norms may constrain the extent to which individual preferences can be translated into outcomes for daughters. Under such conditions, mothers who oppose FGC might face social pressures that limit the correspondence between their attitudes and their daughters’ outcomes. Conversely, if maternal attitudes are more closely reflected in daughter’s FGC outcomes, this pattern may be consistent with mothers having greater scope to influence on outcomes within families and communities. Examining how the correspondence between maternal attitudes and daughter outcomes varies across social contexts may therefore provide insight into the social conditions under which FGC is reproduced across generations. If women’s preferences are substantially constrained in high-prevalence contexts, the correspondence between maternal attitudes and daughter outcomes would be expected to be weaker in regions where the practice is most prevalent. By contrast, it may be that mother preferences are more strongly reflected in daughter FGC outcomes in societies in which traditionalist cultural practices are more common. Clarifying how decision-making operates within families and communities is therefore an important step toward developing empirically grounded interpretations of how cultural practices such as FGC are reproduced across generations.

## The present study

The present study examines whether maternal attitudes toward FGC are associated with daughters’ FGC outcomes, and whether this association varies across contexts in which the practice is more or less prevalent.Using multilevel logistic regression models, we examine whether this association varies as a function of regional prevalence of the practice, controlling for daughter age, mother education, and household wealth. By comparing patterns across regions with different FGC prevalence levels, the analysis examines the social conditions under which maternal attitudes are more or less closely associated with daughter FGC outcomes. Although attitude-outcome correspondence does not directly measure maternal agency, autonomy, or empowerment, variation in this correspondence across contexts may provide evidence relevant to theories concerning women’s decision-making and the role of social and normative constraints in the intergenerational reproduction of FGC.

## METHOD

### Study Design and Data Source

Data were drawn from two independent multinational datasets collected as part of the United Nations Children’s Fund (UNICEF) Multiple Indicator Cluster Surveys^25^. MICS is a large-scale survey program that has collected standardized data on the health and well-being of women and children across more than 120 countries for over three decades. The present analyses use data from the two most recently completed rounds, MICS 5 (2013–2016) and MICS 6 (2017–2022).

MICS data are de-identified, publicly available, and organized by country. The present study used two files from each country: the “FG” file, which contained daughter FGC data, and the “WM” file, which contained the remaining study variables. Data preparation followed MICS guidelines for the merging and management of data files^26^. For each country, the relevant variables from the FG and WM files were merged, after which the country-specific datasets were combined for the MICS 5 and MICS 6 datasets, respectively.

Data were obtained from adult women who self-identified as mothers. Interviews were conducted by trained interviewers in private or semi-private one-to-one settings using standardized questionnaires. Table 1 lists the countries for which FGC data was available and collected, the number of regions per country, number of participants per country, and per-country FGC prevalence rates. Regions correspond to the first administrative subnational level used in MICS surveys^27^. In our multilevel models, daughters constituted Level 1, regions nested within countries constituted Level 2, and countries constituted Level 3 clustering units.

**Table 1.** Per Country Participant, Region, and Prevalence Information.

| **Table 1**<br>**Per Country Participant, Region, and Prevalence Information.** |  |  |  |
| --- | --- | --- | --- |
| Country | Regions per country | Cases per country | Country-level FGC prevalence<sup>^</sup> |
| **MICS 5** |  |  |  |
| Benin | 12 | 12,907 | .23 |
| Cote d' Ivoire | 11 | 25,996 | .52 |
| Guinea | 8 | 11,659 | .99 |
| Guinea-Bissau | 9 | 11,687 | .49 |
| Kenya | 3 | 3,079 | .03 |
| Mali | 8 | 18,748 | 1.00 |
| Mauritania | 13 | 16,482 | .75 |
| Nigeria | 37 | 22,279 | .43 |
| Senegal (Dakar City) | 1 | 7,599 | .18 |
| Total | 102 | 130,436 | .58 |
| **MICS 6** |  |  |  |
| C. African Rep. | 7 | 19,158 | .34 |
| Gambia | 8 | 16,393 | .74 |
| Ghana | 10 | 11,274 | .12 |
| Guinea-Bissau | 9 | 22,288 | .52 |
| Iraq | 18 | 12,568 | .11 |
| Nigeria | 37 | 28,789 | .34 |
| Sierra Leon | 4 | 18,249 | .97 |
| Togo | 7 | 6,046 | .08 |
| Total | 100 | 134,765 | .45 |
| *Note.* ^Percentage of women that reported being circumcised. |  |  |  |

For the MICS 5 dataset, data were obtained from 130,436 respondents from 102 regions across nine African countries. Of these respondents, 97,872 (75%) had complete data on all variables included in the multilevel models and were retained for analysis. For the MICS 6 dataset, data were obtained from 134,765 respondents from 100 regions across one Asian and seven African countries. Of these respondents, 78,853 (59%) had complete data on all study variables and were included in the analytic sample. A substantial proportion of missing daughter FGC status reflects the age-based routing of the MICS questionnaire, in which daughter FGC status was generally not collected for daughters aged 15 years and older. Among daughters within the age range for which FGC status was collected (ages 0-14 years), missing daughter FGC status was uncommon in both survey waves. We compared cases with observed versus missing daughter FGC status on study characteristics (Supplementary Table S1). Aside from the expected difference in daughter age, the observed and missing cases groups showed only small differences in maternal education, household wealth, maternal FGC attitudes, and regional FGC prevalence, suggesting that missing daughter FGC status was unlikely to introduce substantial systematic differences in the analytic samples.

The study was deemed exempt by the Washington State University Institutional Review Board [IRB#20245]. All MICS data were de-identified prior to obtaining it and accessed for the purposes of the present study on 10 May 2023.

### Measures

#### Daughter FGC status

A single dichotomous question was asked of each participant’s mother: “Has she [daughter] been circumcised?” Valid response options for both questions were, *yes* = 1, *no* = 2. For the present study, responses were re-coded so that 0 = uncircumcised and 1 = circumcised.

#### Mother Attitude about circumcision

A single item asked mothers: “Should the practice [of FGC] be continued or discontinued”. A three-point continuous categorical variable was generated based on the valid response options: *Continued* = 1, *Depends or Don’t Know* = 2, and *Discontinued* = 3. High scores reflect a negative attitude toward FGC.

#### Daughter age

Mothers reported age of daughters in years.

#### Mother education

This was assessed with a single item: “What is the highest level and grade or year of school you have attended?” Response options included: *without formal education* = 1; *primary education* (grade one to five) = 2; *secondary education* (grade six to ten) = 3; *higher secondary education and above* (grade 11, 12 and above) = 4.

#### Household wealth

The MICS Wealth Quintile Index assigned households in the entire MICS dataset for each country. They were generated using principal components analysis based on these factors: ownership of household goods and amenities, persons per sleeping room; type of floor, roof, wall, cooking fuel, and sanitary facility; and source of drinking water.

#### Region-level FGC prevalence

This was calculated as the percentage of women in the region sample who answered *yes* to the question, “Have you been circumcised?”

### Statistical Analyses

Multilevel logistic regression modeling was conducted to account for the nesting of participants within regions and countries. Primary analyses explored whether region-level FGC prevalence moderated the association strength between mother FGC attitudes and daughter FGC status. Models included daughter age, maternal education, and household wealth quintile as covariates to account for demographic and socioeconomic differences that may be associated with daughter FGC. Maternal FGC attitudes, region-level FGC prevalence, and their interaction were included as the primary predictors. Country and region-nested-within-country were modeled as random intercepts to account for geographic clustering and country-level heterogeneity. The multilevel models accounted for the hierarchical geographic structure of the data but did not incorporate MICS sampling weights, sampling strata, or primary sampling units as survey-design parameters. Analyses were conducted using *Statistical Package for Social Sciences*, Version 31.

## RESULTS

### Descriptive statistics and regional clustering

Frequencies and descriptive statistics for all study variables are presented in Table 2. Three-level logistic regression models were estimated to assess the degree of clustering in daughter FGC outcomes among individuals nested within regions and countries (Table 3). For the MICS 5 dataset, the estimated country-level variance component was 2.315 (*p* = .065), whereas the region-within-country variance component was 0.809 (p < .001). The corresponding overall adjusted ICC was 0.401, indicating substantial clustering in daughter FGC outcomes across the geographic levels. For the MICS 6 dataset, the estimated country-level variance component was 2.301 *(p* = .106), and the region-within-country variance component was 1.923 (p < .001), with an overall adjusted ICC of 0.365 (p < .001). Thus, approximately 37-40% of the variance in daughter FGC outcomes was attributable to geographic clustering in the two samples. These substantial region-within-country variance and overall ICCs in both datasets support the use of a three-level model to account for the nested structure of the data.

**Table 2.** Descriptive statistics for key study variables across survey waves.

|  | MICS 5 |  |  |  |  | MICS 6 |  |  |  |  |
| --- | --- | --- | --- | --- | --- | --- | --- | --- | --- | --- |
| Variable | N | Mean | SD | Min | Max | N | Mean | SD | Min | Max |
| Daughter FGC status | 100,285 | 0.31 | 0.47 | 0 | 1 | 88,497 | 0.14 | 0.34 | 0 | 1 |
| Daughter age | 129,963 | 9.35 | 7.13 | 0 | 39 | 133,959 | 9.78 | 7.28 | 0 | 40 |
| Mother education | 127,454 | 1.69 | 0.91 | 1 | 4 | 131,502 | 1.93 | 1.05 | 1 | 4 |
| Maternal FGC attitude | 130,214 | 2.25 | 0.92 | 1 | 3 | 125,158 | 2.39 | 0.87 | 1 | 3 |
| Household wealth quintile | 130,436 | 2.91 | 1.39 | 1 | 5 | 134,765 | 2.85 | 1.41 | 1 | 5 |
| Region FGC prevalence | 130,436 | 0.58 | 0.34 | 0.00 | 1.00 | 134,765 | 0.45 | 0.35 | 0.00 | 0.99 |

*Notes.* Maternal FGC attitude coded 1 = Continue, 2 = Don't know, 3 = Discontinue. Means for dichotomous variables represent proportions. Region FGC prevalence is the proportion of circumcised women in each region.

**Table 3.**
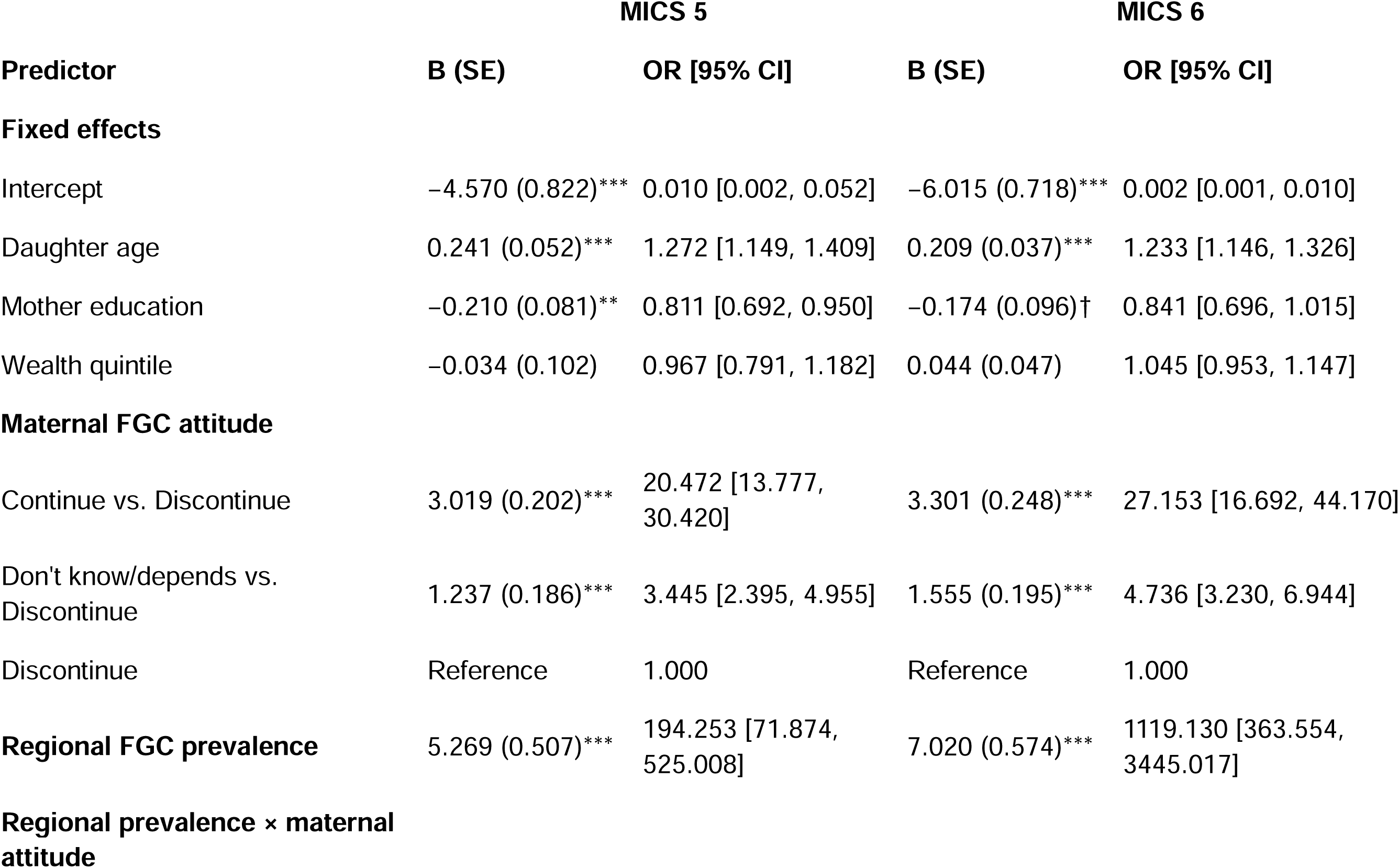
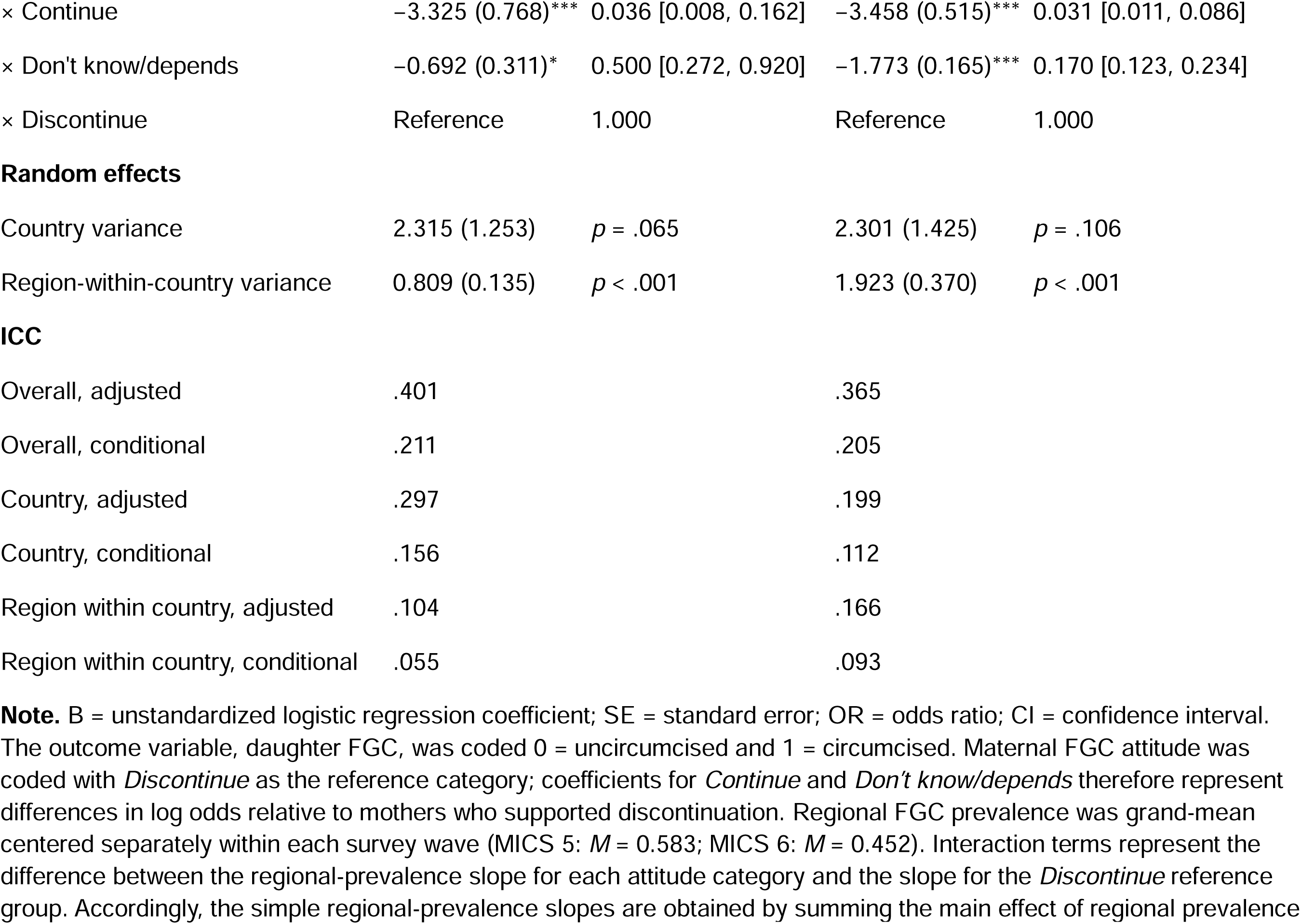
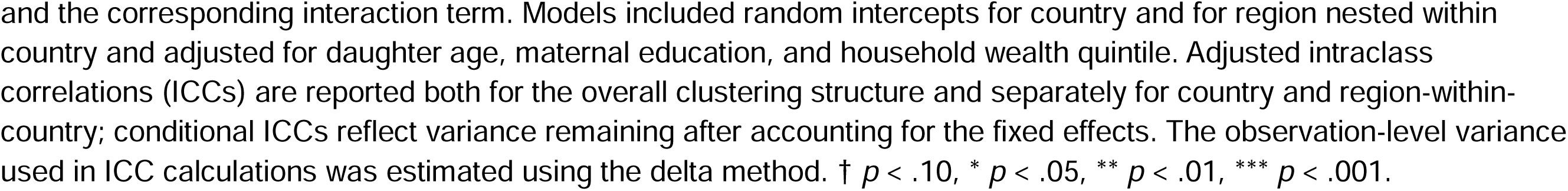
Multilevel Logistic Regression Predicting Daughter FGC as a Function of Maternal FGC Attitudes and Regional FGC Prevalence.

### Multilevel regression models

Multilevel logistic regression models were estimated separately for the MICS 5 and MICS 6 datasets. The dependent variable was daughter circumcision status (0 = uncircumcised, 1 = circumcised). Models included daughter age, maternal education, household wealth quintile, and maternal FGC attitudes. Country and region-nested-within-country were modeled as random intercepts, yielding a three-level structure in which daughters were nested within regions and regions were nested within countries.

Maternal attitudes toward FGC were strongly associated with daughter circumcision outcomes in both survey waves. Relative to mothers who supported FGC discontinuation, mothers endorsing continuation had substantially higher odds of having circumcised daughters (MICS 5: OR = 20.47, 95% CI [13.78, 30.42], p < .001; MICS 6: OR = 27.15, 95% CI [16.69, 44.17], p < .001). Similarly, mothers who reported that they did not know or that the decision depended on circumstances had higher odds of having circumcised daughters (MICS 5: OR = 3.45, 95% CI [2.40, 4.96], p < .001; MICS 6: OR = 4.74, 95% CI [3.23, 6.94], p < .001). These coefficients represent differences between maternal attitude groups at the mean level of regional FGC prevalence, because regional prevalence was grand-mean centered within each survey wave.

Regional prevalence was positively associated with daughter FGC among mothers who supported discontinuation (MICS 5: B = 5.269, OR = 194.25, 95% CI [71.87, 525.01], *p* < .001; MICS 6: B = 7.020, OR = 1,119.13, 95% CI [363.55, 3,445.02], *p* < .001). However, the interaction between regional prevalence and maternal FGC attitudes indicated that this association was significantly weaker among mothers supporting continuation (MICS 5: B = -3.325, OR = 0.036, 95% CI [0.008, 0.162], *p* < .001; MICS 6: B = -3.458, OR = 0.031, 95% CI [0.011, 0.086], *p* < .001), and among mothers reporting uncertainty (MICS 5: B = -0.692, OR = 0.500, 95% CI [0.272, 0.920], p = .026; MICS 6: B = -1.773, OR = 0.170, 95% CI [0.123, 0.234], *p* < .001). Simple-slope analyses showed that regional prevalence was positively associated with daughter FGC for all three attitude groups, but the slope was weakest among mothers supporting FGC continuation, intermediate among mothers reporting uncertainty, and strongest among mothers supporting discontinuation. This pattern was observed for both MICS 5 and MICS 6 (Table 3).

### Cross-level interaction effects

Because mothers supporting FGC discontinuation constituted the reference group, the negative interaction coefficients indicate that the positive association between regional FGC prevalence and daughter FGC was weaker among mothers supporting continuation or expressing uncertainty than among mothers supporting discontinuation. Simple-slope analyses confirmed that regional prevalence was positively associated with daughter FGC in all three maternal attitude groups, but the association was weakest among mothers supporting continuation, intermediate among mothers expressing uncertainty, and strongest among mothers supporting discontinuation. This pattern was replicated in both MICS 5 and MICS 6 (Table 3; Figures 1 and 2).

**Figure 1.**
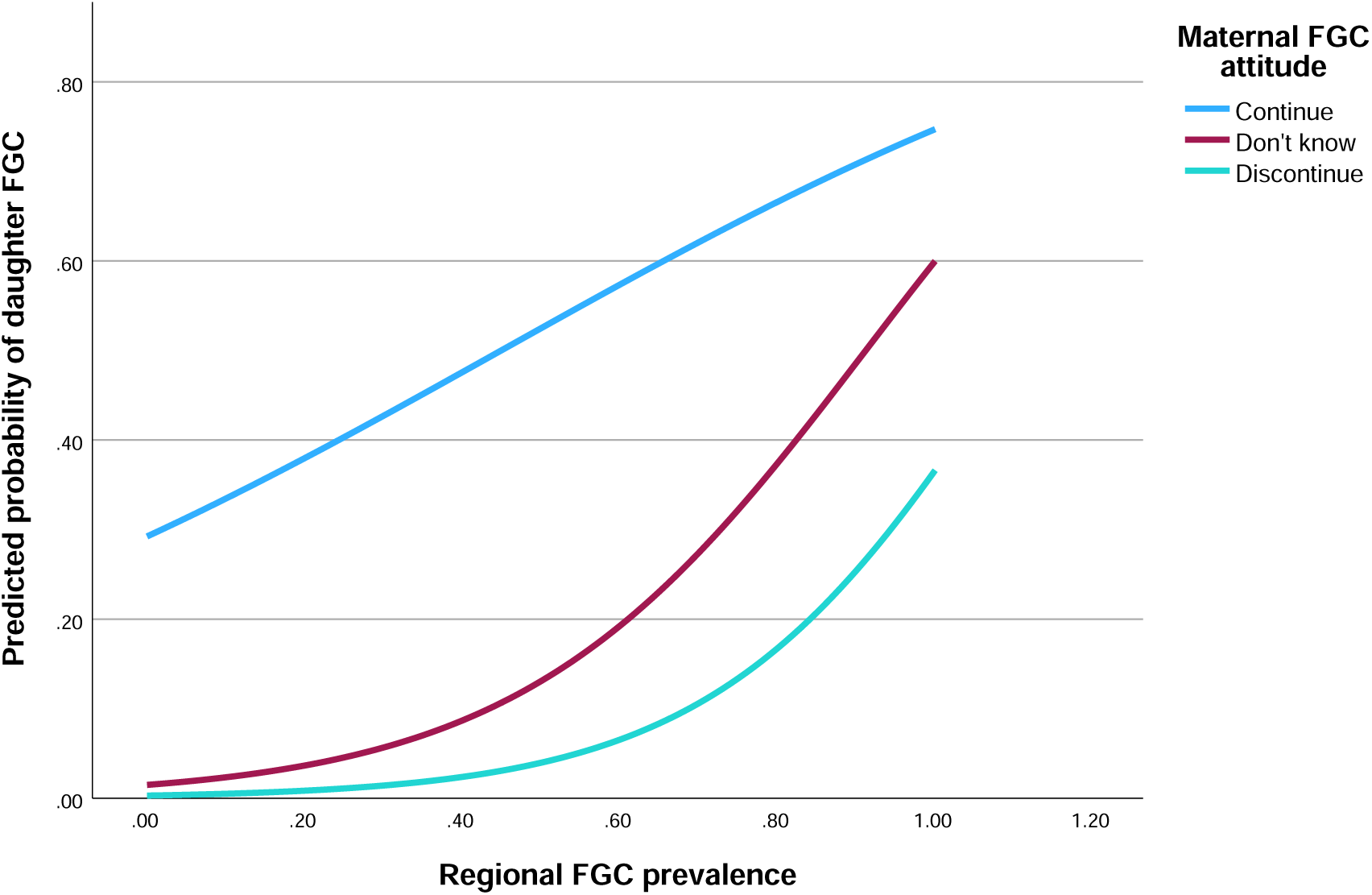
Predicted probability of daughter FGC by region FGC prevalence and maternal attitudes (MICS 5).

**Figure 2.**
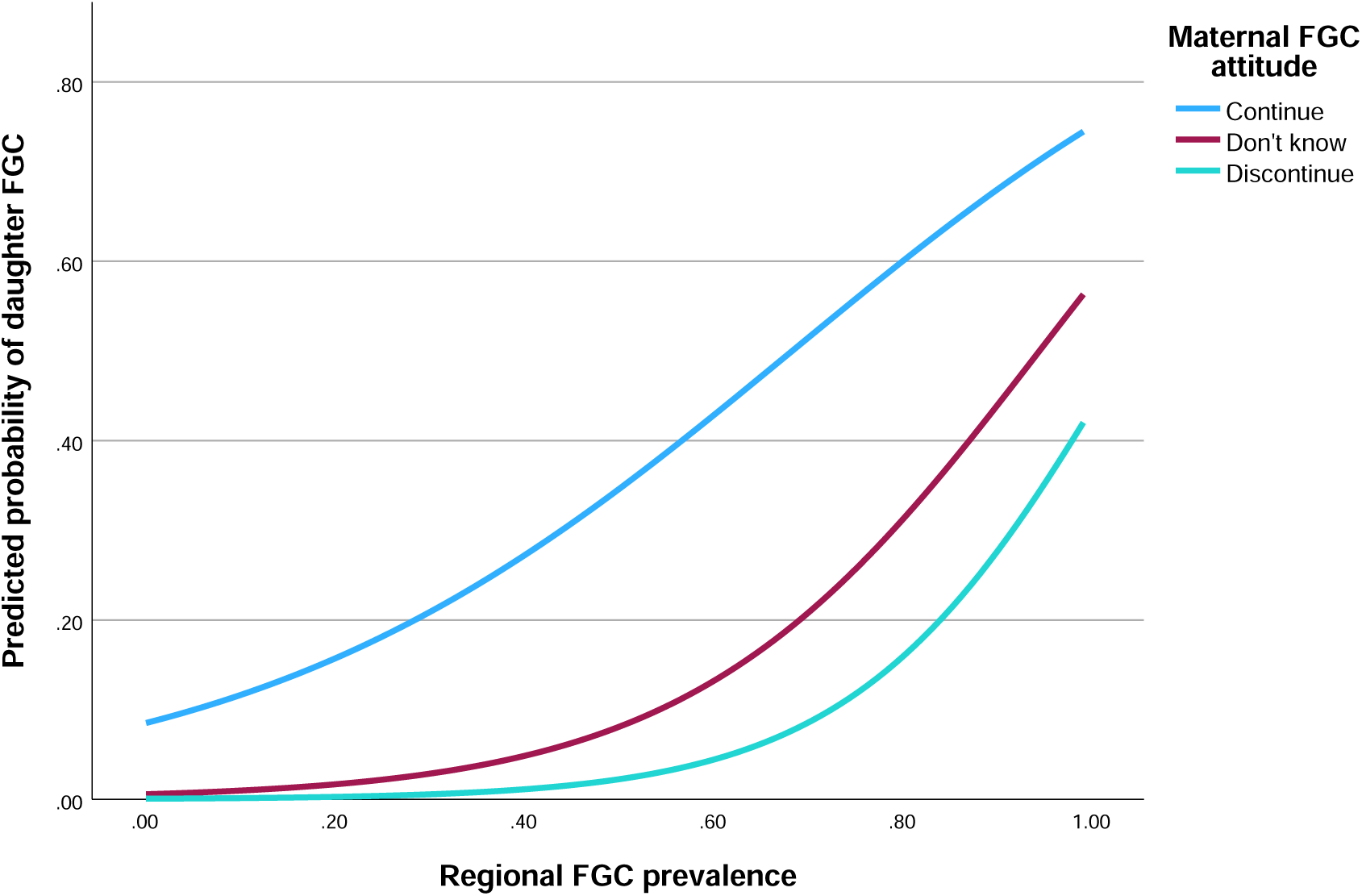
Predicted probability of daughter FGC by region FGC prevalence and maternal attitudes (MICS 6).

### Marginal effects and predicted probabilities

To facilitate interpretation of the interaction effects, predicted probabilities of daughter circumcision were calculated across the observed range of regional FGC prevalence for each maternal attitude category. Across both survey waves, predicted probabilities increased as regional FGC prevalence increased within all three maternal attitude groups (Figures 1 and 2). However, the rate of increase differed by maternal attitude. The probability of daughter FGC increased most rapidly among mothers who supported discontinuation, increased at an intermediate rate among mothers expressing uncertainty, and increased most slowly among mothers supporting continuation. Consequently, differences in predicted probabilities between maternal attitude groups were largest at lower levels of regional FGC prevalence and diminished as regional prevalence increased. This pattern was observed in both MICS 5 and MICS 6 and is consistent with the simple-slope analyses reported in Table 3. Thus, the association between maternal attitudes and daughter FGC was stronger in lower-prevalence contexts, a pattern consistent with the cultural repression hypothesis.

### Sensitivity Analyses

Because daughter FGC status may be age-dependent, we conducted sensitivity analyses restricting the sample to daughters aged ≥10 years. This threshold was supported by the observed distribution of age at FGC in the MICS data, with 99.5% of daughters with reported age-at-FGC information having undergone FGC by age 10. The results were consistent with those of the primary analyses in both MICS 5 and MICS 6 (Supplementary Table S2). Thus, the interaction pattern was not dependent on the inclusion of younger daughters for whom FGC status may not yet have been fully determined.

## DISCUSSION

The present study examined whether maternal attitudes toward female genital cutting (FGC) are associated with daughters’ FGC outcomes and whether the strength of this association varies across societies in which the practice occurs. Across two large multinational datasets, the correspondence between maternal attitudes toward FGC and daughters’ FGC status was stronger in lower-prevalence regions.This pattern is consistent with the cultural repression hypothesis 6, ^28^. This hypothesis suggests that in regions where FGC is highly prevalent and strongly normative, social expectations and pressures may constrain the extent to which individual attitudes of women are reflected in outcomes. That is, a mother’s opposition to FGC would be expected to have a weaker correspondence with whether her daughter undergoes FGC in high-prevalence contexts. The consistent pattern observed across MICS 5 and MICS 6 is compatible with this interpretation.

These findings are relevant to an understanding of the social dynamics through which FGC is maintained and reproduced across generations. Previous ethnographic and anthropological research has emphasized that the meanings and social functions of FGC vary across communities. By some accounts, FGC is embedded in women-centered institutions, initiation practices, systems of social status and solidarity, and women play important roles in maintaining and regulating the practice^14, 17, 23, 29^. Other researchers, drawing on social convention and social norms perspectives, emphasize how factors such as marriageability, social acceptance, and sanctions for deviating from community expectations may sustain FGC even as attitudes toward the practice change^23^.

One possible interpretation of the present findings is that, where FGC is more widely practiced, normative contexts may constrain the extent to which differences in maternal attitudes get reflected in outcomes for daughters. This is consistent with the cultural disempowerment hypothesis. According to this hypothesis, FGC is reflective of a broader pattern of social norms and expectations in which women’s views on culturally informed practices are less closely reflected in outcomes.

The present findings may help contextualize findings from previous research showing that daughter FGC was positively associated with maternal happiness, primarily in countries where FGC was less prevalent^30^. At face value, this association appears contradictory to cultural repression given daughter FGC was associated with a more positive maternal emotional outcome. The present findings suggest a different interpretation of this pattern. Specifically, if contexts in which FGC is less common provide women greater scope to have their attitudes and preferences reflected in decisions, then the positive association between daughter FGC and maternal happiness observed by Strand et al. (2024)^30^ may reflect greater agency or empowerment in these settings. This interpretation is consistent with evidence that women’s life satisfaction is higher in environments characterized by less restrictive gender norms^31, 32^.

### Limitations

Limitations of the present study include the following. First, the correspondence between maternal attitudes and daughter FGC status should not be interpreted as a direct measure of maternal agency, autonomy, empowerment, or decision-making authority. Maternal attitudes and daughter FGC outcomes may both reflect family expectations, community norms, social pressures, economic circumstances, or other factors not measured in the present study. Thus, greater attitude–outcome correspondence does not necessarily indicate greater autonomy or unconstrained choice. Second, the analyses relied on cross-sectional survey data and therefore cannot establish causal relationships or determine whether regional prevalence produces differences in attitude–outcome correspondence. Third, some key variables were measured using single survey items, which may introduce measurement error. Fourth, reporting biases may occur in settings where FGC is legally restricted or socially stigmatized, potentially affecting reported attitudes and FGC status. Finally, the countries included in the analysis represent only those participating in the relevant MICS survey rounds, and the findings may not generalize to all societies in which FGC is practiced.

## Conclusion

Across two multinational MICS datasets, the association between maternal FGC attitudes and daughter FGC outcomes was stronger in regions where FGC was less prevalent and weaker in regions where FGC was more prevalent. Specifically, differences in predicted daughter FGC probabilities across maternal attitude groups were larger in lower-prevalence regions and diminished as regional prevalence increased. This pattern is consistent with the possibility that stronger social and normative contexts surrounding FGC may constrain the extent to which maternal attitudes are reflected in daughters’ outcomes. These findings are consistent with notions that higher rates of the intergenerational transmission of FGC may align with social factors that interfere with women’s influence on culturally-informed decisions such as those concerning FGC.

## Supporting information

Syntax Supplement

## Data Availability

All data are de-identified, organized by country, and publicly available at https://mics.unicef.org/surveys.

