## Supplemental Table S1 for "Female genital cutting and maternal attitudes about it: Testing a cultural disempowerment hypothesis across two large multi-country samples"

**Supplementary Table S1. Comparison of Observations With Observed Versus Missing Daughter FGC Status**

|  | **MICS 5** | | | **MICS 6** | | |
| --- | --- | --- | --- | --- | --- | --- |
| **Variable** | **Observed FGC** | **Missing FGC** | **SMD** | **Observed FGC** | **Missing FGC** | **SMD** |
| **N** | 100,285 | 30,151 | — | 100,564 | 34,201 | — |
| **Daughter age, M (SD)** | 6.23 (4.19) | 19.89 (4.39) | -3.23 | 6.33 (4.20) | 19.85 (4.43) | -3.22 |
| **Mother education, M (SD)** | 1.72 (0.93) | 1.61 (0.84) | 0.12 | 1.97 (1.07) | 1.83 (1.02) | 0.13 |
| **Household wealth quintile, M (SD)** | 2.89 (1.39) | 2.97 (1.38) | -0.06 | 2.84 (1.41) | 2.91 (1.42) | -0.05 |
| **Combined wealth index, M (SD)** | -0.05 (0.94) | 0.01 (0.94) | -0.06 | -0.01 (0.95) | 0.04 (0.97) | -0.05 |
| **Maternal FGC attitude, M (SD)** | 2.23 (0.93) | 2.33 (0.90) | -0.11 | 2.38 (0.87) | 2.41 (0.87) | -0.03 |
| **Regional FGC prevalence, M (SD)** | 0.591 (0.344) | 0.554 (0.345) | 0.11 | 0.456 (0.353) | 0.439 (0.355) | 0.05 |

**Note.** M = mean; SD = standard deviation; SMD = standardized mean difference. Daughter FGC status was coded 0 = uncircumcised and 1 = circumcised. Missing daughter FGC status includes observations for which the daughter FGC variable was not available; a substantial proportion of these observations reflects age-based survey routing rather than item nonresponse. In MICS 5, daughter FGC status was observed for 100,285 observations and missing for 30,151; in MICS 6, daughter FGC status was observed for 100,564 observations and missing for 34,201. The large standardized differences in daughter age reflect the age-based eligibility structure of the daughter FGC module and should not be interpreted as evidence of differential attrition. SMDs are calculated as the difference between group means divided by the pooled standard deviation; absolute values below approximately 0.10 are generally considered small, with values around 0.10–0.20 indicating at most modest differences.
