## Supplemental Table S2 for "Female genital cutting and maternal attitudes about it: Testing a cultural disempowerment hypothesis across two large multi-country samples"

| **Supplementary Table S2. Multilevel Logistic Regression Predicting Daughter FGC Among Daughters Aged ≥10 Years** | | | | |
| --- | --- | --- | --- | --- |
|  | **MICS 5** | | **MICS 6** | |
| **Predictor** | **B (SE)** | **OR [95% CI]** | **B (SE)** | **OR [95% CI]** |
| **Fixed effects** |  |  |  |  |
| Intercept | −2.790 (.512)*** | .061 [.023, .168] | −4.848 (1.001)*** | .008 [.001, .056] |
| Daughter age | .108 (.023)*** | 1.114 [1.065, 1.165] | .129 (.058)* | 1.138 [1.017, 1.274] |
| Mother education | −.267 (.111)* | .766 [.616, .951] | −.198 (.121)† | .820 [.648, 1.039] |
| Wealth quintile | −.047 (.121) | .954 [.753, 1.209] | .022 (.068) | 1.022 [.894, 1.167] |
| **Maternal FGC attitude** |  |  |  |  |
| Continue vs. Discontinue | 2.981 (.098)*** | 19.716 [16.262, 23.902] | 2.990 (.232)*** | 19.887 [12.631, 31.312] |
| Don't know/depends vs. Discontinue | 1.183 (.233)*** | 3.263 [2.065, 5.156] | 1.408 (.216)*** | 4.088 [2.677, 6.241] |
| Discontinue | Reference | — | Reference | — |
| **Regional FGC prevalence** |  |  |  |  |
| Regional prevalence | 4.530 (.299)*** | 92.770 [51.615, 166.740] | 6.336 (.424)*** | 564.436 [245.809, 1,296.079] |
| Prevalence × Continue | −2.057 (.539)*** | .128 [.044, .368] | −2.465 (.903)** | .085 [.014, .498] |
| Prevalence × Don't know/depends | −.258 (.211) | .772 [.510, 1.169] | −1.170 (.294)*** | .310 [.174, .553] |
| Prevalence × Discontinue | Reference | — | Reference | — |
| **Simple slopes of regional prevalence** |  |  |  |  |
| Continue | 2.473 (.583)* | 11.86 [3.78, 37.20] | 3.871 (.672)* | 47.99 [12.87, 179.01] |
| Don't know/depends | 4.272 (.402)* | 71.67 [32.58, 157.65] | 5.166 (.508)* | 175.21 [64.72, 474.35] |
| Discontinue | 4.530 (.299)* | 92.76 [51.61, 166.72] | 6.336 (.424)* | 564.53 [245.86, 1,296.28] |
| **Random effects** |  |  |  |  |
| Country variance | 1.690 (.914) | — | 1.679 (1.006) | — |
| Region within country variance | .475 (.094)*** | — | 1.025 (.225)*** | — |
| **Intraclass correlations** |  |  |  |  |
| Overall, adjusted | .345 | — | .327 | — |
| Overall, conditional | .179 | — | .173 | — |
| Country, adjusted | .269 | — | .203 | — |
| Country, conditional | .140 | — | .107 | — |
| Region within country, adjusted | .076 | — | .124 | — |
| Region within country, conditional | .039 | — | .065 | — |

**Note.** B = unstandardized logistic regression coefficient; SE = standard error; OR = odds ratio; CI = confidence interval. Daughter FGC was coded 0 = uncircumcised and 1 = circumcised. Maternal FGC attitude was coded with Discontinue as the reference category. Regional FGC prevalence was grand-mean centered separately within each survey wave. The interaction terms represent differences between the regional-prevalence slope for each attitude category and the slope for the Discontinue reference group. Simple slopes represent the estimated association between regional FGC prevalence and daughter FGC within each maternal attitude category and were calculated by summing the regional-prevalence main effect and the corresponding interaction coefficient. Models were restricted to daughters aged ≥10 years and adjusted for daughter age, maternal education, and household wealth quintile. Models included random intercepts for country and region nested within country. Intraclass correlations (ICCs) are reported for the overall clustering structure and separately for country and region-within-country; conditional ICCs reflect variance remaining after accounting for the fixed effects. Observation-level variance was estimated using the delta method. **p* < .01, ***p* < .001.
