## Supplementary material for "Female genital cutting and maternal attitudes about it: Testing a cultural disempowerment hypothesis across two large multi-country samples": Syntax Supplement

| **Supplementary Materials: SPSS Syntax for output reported in Table 3.** | |
| --- | --- |
| **Syntax** | GENLINMIXED /DATA_STRUCTURE SUBJECTS=CC*HH7_unique /FIELDS TARGET=FG15r TRIALS=NONE OFFSET=NONE /TARGET_OPTIONS REFERENCE=0 DISTRIBUTION=BINOMIAL LINK=LOGIT /FIXED EFFECTS=FG13 welevel windex5 FG22r Region_Prev_C Region_Prev_C*FG22r USE_INTERCEPT=TRUE /RANDOM USE_INTERCEPT=TRUE SUBJECTS=CC COVARIANCE_TYPE=VARIANCE_COMPONENTS SOLUTION=FALSE /RANDOM USE_INTERCEPT=TRUE SUBJECTS=CC*HH7_unique COVARIANCE_TYPE=VARIANCE_COMPONENTS SOLUTION=FALSE /BUILD_OPTIONS TARGET_CATEGORY_ORDER=ASCENDING INPUTS_CATEGORY_ORDER=ASCENDING HCONVERGE=0.00000001(RELATIVE) MAX_ITERATIONS=100 CONFIDENCE_LEVEL=95 DF_METHOD=RESIDUAL COVB=ROBUST SCORING=0 SINGULAR=0.000000000001 /EMMEANS TABLES=FG22r CONTRAST=NONE /EMMEANS_OPTIONS SCALE=ORIGINAL PADJUST=LSD. |
|  | *Notes*. CC = Country code; HH7_unique = Subnational Region; FG15r = daughter circumcision status; FG13 = Daughter age; welevel = woman (mother) education level; windex5 = household wealth quintile; FG22r = mother attitude about circumcision; Region_Prev_C = Region-level mother circumcision prevalence. |
